# Characteristics for Machine Learning Detection of Large Vessel Occlusion on Computed Tomography Angiography

**DOI:** 10.1101/2023.02.13.23285882

**Authors:** Sneha Lingam, Lucas W. Remedios, Samuel W. Remedios, Bennett A. Landman, Larry T. Davis, Stephen W. Clark

**Affiliations:** School of Medicine, Vanderbilt University, Nashville, TN, USA; Medical-image Analysis and Statistical Interpretation Lab, Vanderbilt Institute for Surgery and Engineering, Nashville, TN, USA; Department of Computer Science, Vanderbilt University, Nashville, TN, USA; Department of Computer Science, Johns Hopkins University, Baltimore, MD, USA; Department of Radiology and Imaging Sciences, National Institutes of Health, Bethesda, MD, USA; Department of Electrical Engineering, Vanderbilt University, Nashville, TN, USA; Department of Radiology and Radiological Sciences, Vanderbilt University Medical Center, Nashville, TN, USA; Department of Neurology, Vanderbilt University Medical Center, Nashville, TN, USA; Sir Galahad Labs, Nashville, TN; Epiphany Biosciences, San Francisco, CA

**Author notes:** Corresponding Authors: Taylor Davis, Stephen Clark.

**Keywords:** stroke, machine learning, computed tomography angiography, artificial intelligence, embolic stroke

## Abstract

Detection of large vessel occlusion (LVO) using machine learning on computed tomography angiography (CTA) may help stroke triage, yet applicability across varied patient and image characteristics has not been examined. The study will examine which characteristics are important when using a convolutional neural network to identify LVO on CTA. A retrospective cohort study (November 2017–May 2019) at a comprehensive stroke center evaluated 677 stroke-alerted patients with an LVO of the internal carotid artery, M1, or M2 (n=150) and a matching number without LVO were included. An Inception module-based network was trained for binary classification of LVO presence. Results were examined by LVO location, window settings, non-LVO findings, demographics, risk factors, presentation status and times, interventions, and outcomes. Three hundred patients were included (48% women; median age 65). Mean±95% CI for cross-validation test and external validation, respectively, are area under precision-recall curve 0.871±0.094 and 0.742±0.018 and area under receiver operating characteristic curve 0.920±0.051 and 0.852±0.004. 145 true positive (TP), 5 false negative (FN), 39 false positive (FP), and 111 true negative (TN) patients were identified. Significant comparisons (P<0.05) identified: lower window settings for misclassifications, smoking history for all FN versus 33% TP (P=0.005), and tissue plasminogen activator treatment for 41% FP versus 20% TN (P=0.017). Our LVO detection tool had high performance across patient characteristics with few exceptions. FP had pathology warranting detection, including distal occlusions. Lower window settings among misclassifications highlight the need for image quality when using machine learning for decision support.

## Introduction

Patients with acute ischemic stroke due to LVO are at high risk for poor outcomes and benefit from early identification and reperfusion.^1–7^ Automated detection of LVO using machine learning on the increasingly available imaging modality of CTA may help stroke triage.^8–11^ Several studies have successfully demonstrated this application with high performance, yet applicability across varied patient and image characteristics has not been examined.^12–17^

Any machine learning algorithm is biased by the population included and data that it is trained on, which prompts the need to critically evaluate performance of LVO detection algorithms in the context of patient and image characteristics.^18,19^ Many factors are known to cause variation in imaging appearance of vessels and LVO, including LVO location, age-related vascular changes, risk factors such as smoking and hypertension, and time from stroke onset to imaging.^20–23^ CTA-specific properties such as contrast enhancement also vary on a patient-to-patient basis.^24^ It is also worth investigating algorithm performance relative to interventions and outcomes, since these can be associated with stroke features and severity reflected in CTA.^25,26^ Algorithm evaluation in the context of such factors can identify any that are associated with poor performance and may require closer attention when using an automated LVO detection tool for clinical decision support.

The aim of this study is to determine what patient and image characteristics are important when using a convolutional neural network (CNN) to identify potential LVO from CTA. To this end, a CNN was trained on the task of LVO detection from CTA, and performance was subsequently examined in the context of patient demographics, stroke risk factors, clinical status on arrival, interventions, outcomes, time intervals during triage and intervention, LVO location, image window settings, and other imaging findings.

In this retrospective study, we aimed to determine which patient and imaging characteristics are important when training a convolutional neural net to identify LVO on CTA.

## Methods

### Patients and Data

This retrospective cohort study was approved by the Vanderbilt University institutional review board (#191100). Among 677 stroke-alerted patients at a comprehensive stroke center during the period November 2017–May 2019, 150 with anterior circulation LVO and 150 without LVO were included, with selection process detailed in Figure 1. Only the first stroke was considered for patients who had multiple during the timeframe. LVO is defined as occlusion of the internal carotid artery (ICA) or the middle cerebral artery’s first (M1) or proximal second (M2) segment. LVO labels were per chart review, or if ambiguous in documentation, images were re-examined by author LTD, a board-certified neuroradiologist. Exclusion criteria were patients without head non-contrast computed tomography (NCCT) or CTA available; images that were unsuccessfully transferred to the research server, uninterpretable as far as LVO label, or distorted upon processing; intracranial hemorrhage or implant such as external ventricular drain; and rare pathology such as posterior circulation occlusion, common carotid artery occlusion, intracranial mass, and moyamoya.

**Figure 1:**
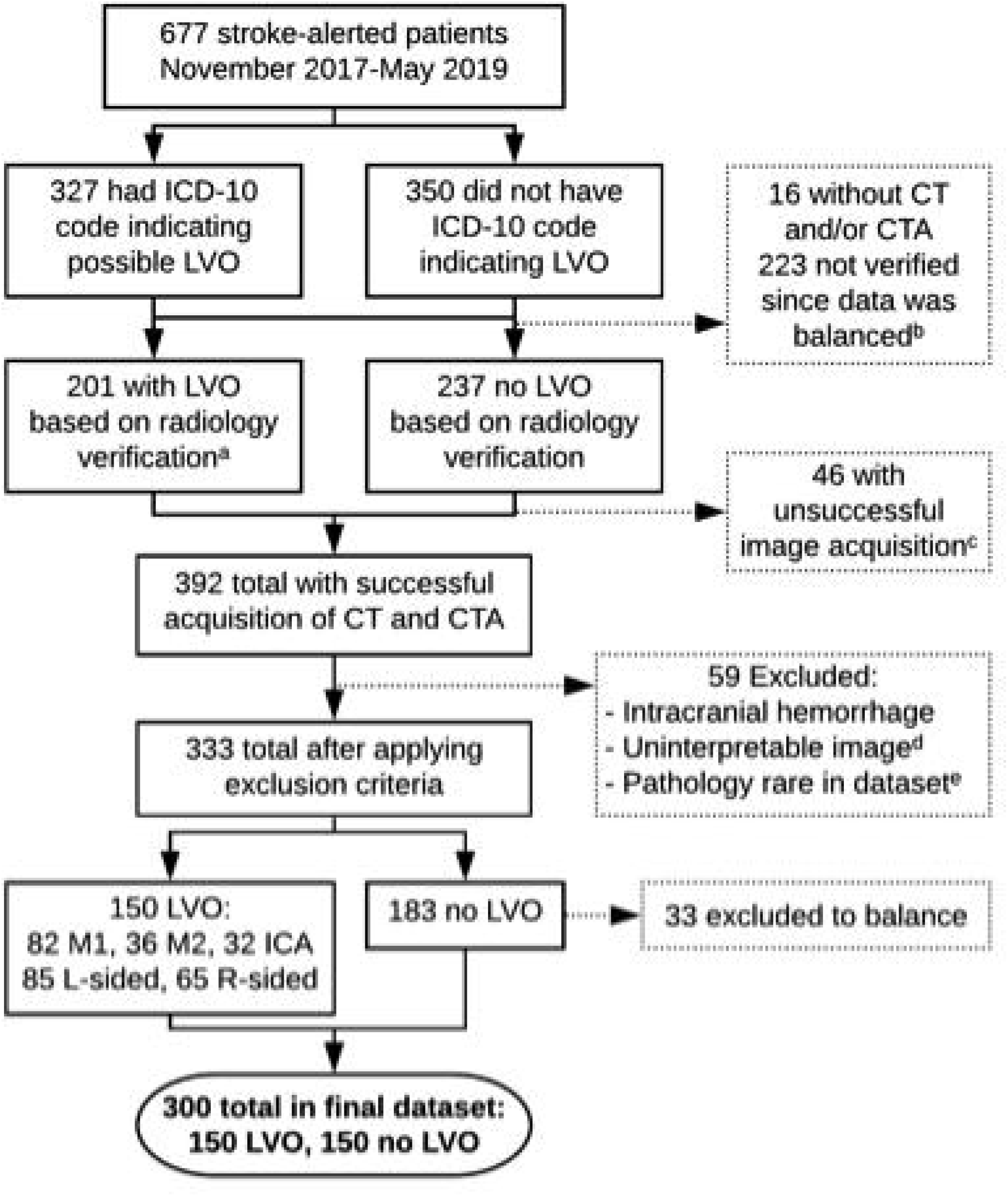
Patient selection flow diagram. (a) Radiology reports were used for verification of LVO or cutoff. In case of ambiguity, images were re-examined by author LTD, a board-certified neuroradiologist. (b) Aimed to have equal numbers of LVO and no LVO examples and stopped verification when data was balanced. All 324 with LVO-related ICD codes and images available were verified, of which 196 had LVO and 128 did not; of other ICD codes, 109 of 114 checked did not have LVO, at which point verification was stopped. (c) Images failed to transfer to our server or were missing slices. (d) Original image was uninterpretable (e.g. due to severe motion degradation) and/or image was distorted upon registration as part of preprocessing. (e) Rare pathology consisted of posterior circulation occlusion (n=12), mass lesion, moyamoya, surgical device such as external ventricular drain, and common carotid artery occlusion.

Stroke alert database was provided by the Vanderbilt Stroke Program. Patient characteristics were acquired from electronic medical records, securely recorded using Research Electronic Data Capture (REDCap), and deidentified and date-shifted prior to downloading for analysis.^27,28^ Characteristics considered include demographics, stroke risk factors, whether patient was transferred, National Institutes of Health Stroke Scale (NIHSS), blood pressure on arrival, interventions, outcomes, timestamps during triage and intervention, and radiology reports. NCCT of the head and thin slice (1mm) CTA of the combined head and neck were acquired in Digital Imaging and Communications in Medicine (DICOM) format from Vanderbilt University Medical Center databases in Agfa IMPAX 6 and ImageVU, deidentified, and stored in the Vanderbilt University Institute of Imaging Science Center for Computational Imaging XNAT database.^29^

### Image Processing

Open-source tools were used for image processing.^30–33^ NCCT and CTA were converted from DICOM to Neuroimaging Informatics Technology Initiative format, CTA were cropped to include head only, and all images were registered to a common template NCCT selected from the dataset. Skull was removed by using a validated brain extraction method on each patient’s NCCT and then applying the mask from NCCT skull removal to the same patient’s CTA.^34^ For each CTA, Hounsfield unit window width and level were adjusted based on visual inspection, a 40mm axial maximum intensity projection image (MIP) to optimally depict anterior circulation was generated, and intensity was normalized to range 0-1. The CNN PhiNet was tested for feasibility of training on this dataset using images at each step of processing, and it was only able to learn effectively after application of manually involved steps of individualized window adjustment and reduction from three to two dimensions with MIP generation.

### Model Training, Evaluation, and Selection

Five deep CNNs were trained for binary classification of LVO or no LVO, using 240 CTA MIPs with 10-fold cross-validation. A balanced set of 60 images was withheld for external validation. The 240 cross-validation images were overall balanced between labels, but split into train, validation, and test sets for each fold did not specify balanced classes. For each fold, 10% (24 images) were used as a test set, and remaining images were split 80:20 for train and validation. Each of the 240 images was included in the cross-validation test set for exactly one fold. PhiNet, EfficientNet-B0, DenseNet-121, ResNet-50, and a network based on the Inception V1 module were implemented with Keras and TensorFlow.^35–42^ The output prediction of LVO presence was a value between 0-1.

For each CNN, all 2400 predictions from the ten folds of cross-validation (train, validation, and test sets) were aggregated, F1 score was computed at every possible discrimination threshold, and the threshold was selected where F1 was highest. The threshold was applied to each prediction to determine a predicted class. The ten trained models from each fold of cross-validation were evaluated separately on 1) the corresponding fold’s cross-validation test set of 24 images and 2) the common external validation set of 60 images. Evaluation metrics were accuracy, sensitivity, precision, specificity, and F1 score at the selected threshold, area under precision-recall curve (AUPRC), and area under receiver operating characteristic curve (ROC-AUC). Separately for cross-validation and external validation sets, each metric was averaged across folds and 95% confidence intervals (CI) computed. The CNN with highest sum of the seven mean metrics for the cross-validation test set was selected. Results are reported solely for the selected CNN.

### Characterization and Error Analysis

Each patient had a single prediction from each of the ten folds when all cross-validation (train, validation, and test sets) and external validation set predictions were aggregated. The ten predictions for each of the 300 patients were averaged. The selected threshold was applied to each patient’s average prediction to determine one predicted class per patient and sort into true positive (TP), false negative (FN), false positive (FP), and true negative (TN) groups. LVO location, window settings, demographics, known history of stroke risk factors, presentation status and times, interventions, and outcomes were compared for TP versus FN and FP versus TN using Fisher’s Exact Test for Count Data for categorical variables and the Wilcoxon Rank Sum Test for continuous variables, at a significance level of P<0.05. Information regarding non-LVO pathology and artifacts for patients in each group was obtained from radiology reports.

## Results

### Patient Characteristics

Among 300 patients, 145 (48%) were women, and median age (interquartile range [IQR]) was 65 (55-76). LVO included 32 (21%) ICA, 82 (55%) M1, and 36 (24%) M2. Patient characteristics by LVO label are detailed in Table 1. Characteristics not reported due to low documentation rates are modified Rankin scale, 24-hour NIHSS, and Alberta Stroke Program Early CT score (ASPECTS).

**Table 1:** Patient characteristics by LVO label

| **Characteristic** | **LVO (n=150)** | **No LVO (n=150)** |
| --- | --- | --- |
| **DEMOGRAPHICS** |  |  |
| Age, median (IQR) | 65 (55-76) | 66 (54-76) |
| Female, n (%) | 66 (44%) | 79 (53%) |
| Race, n (%) |  |  |
| Black | 20 (13%) | 26 (17%) |
| White | 123 (82%) | 119 (79%) |
| Other or Unknown<sup>a</sup> | 7 (5%) | 5 (3%) |
| Ethnicity, n (%) |  |  |
| Hispanic or Latinx | 3 (2%) | 5 (3%) |
| Not Hispanic or Latinx | 141 (94%) | 141 (94%) |
| Unknown | 6 (4%) | 4 (3%) |
| **PAST MEDICAL HISTORY, n (%)** |  |  |
| Hypertension | 95 (63%) | 99 (66%) |
| Diabetes | 33 (22%) | 42 (28%) |
| Hyperlipidemia | 55 (37%) | 53 (35%) |
| Smoking | 53 (35%) | 46 (31%) |
| Coronary Artery Disease | 33 (22%) | 30 (20%) |
| Congestive Heart Failure | 21 (14%) | 15 (10%) |
| Atrial Fibrillation | 42 (28%) | 21 (14%) |
| On Anticoagulation Medication | 29 (19%) | 14 (9%) |
| Prior Stroke or TIA | 29 (19%) | 31 (21%) |
| **STATUS ON ARRIVAL** |  |  |
| Transfer from Outside Hospital, n (%) | 101 (67%) | 57 (38%) |
| Pretreatment NIHSS<sup>b</sup>, median (IQR) | 14 (8-19) | 4 (1-11) |
| Systolic Blood Pressure, median (IQR), mmHg | 155 (140-173) | 156 (137-185) |
| Diastolic Blood Pressure<sup>c</sup>, median (IQR), mmHg | 85 (76-100) | 89 (77-103) |
| **LVO LOCATION** |  |  |
| Left-sided, n (%) | 85 (57%) | N/A |
| Artery, n (%) |  |  |
| ICA | 32 (21%) | N/A |
| M1 | 82 (55%) | N/A |
| M2 | 36 (24%) | N/A |
| **INTERVENTION** |  |  |
| Received tPA, n (% out of all patients) | 59 (39%) | 38 (25%) |
| Received tPA, n (% out of patients with LKW to CT ≤4.5 hours)<sup>d</sup> | 47 (76%) | 30 (48%) |
| Subsequent Thrombectomy, n (%) | 83 (55%) | N/A |

**Table 1: Patient characteristics by LVO label (continued)**
| Characteristic | LVO (n=150) | No LVO (n=150) |
| --- | --- | --- |
| OUTCOMES |  |  |
| Reperfusion, n (% out of patients who had thrombectomy) | 72 (87%) | N/A |
| TICI Score, n (% out of patients who had thrombectomy) |  |  |
| 0 | 10 (12%) | N/A |
| 1 | 1 (1%) | N/A |
| 2a | 0 (0%) | N/A |
| 2b | 25 (30%) | N/A |
| 2c | 13 (16%) | N/A |
| 3 | 34 (41%) | N/A |
| Length of Stay, median (IQR), days | 5 (3-9) | 3 (1-5) |
| Disposition, n (%) |  |  |
| Expired | 19 (13%) | 4 (3%) |
| Home | 43 (29%) | 96 (64%) |
| Rehab Facility | 37 (25%) | 26 (17%) |
| Skilled Nursing Facility | 34 (23%) | 18 (12%) |
| Hospice | 8 (5%) | 2 (1%) |
| Other | 9 (6%) | 4 (3%) |
| TIME INTERVALS, median (IQR), minutes |  |  |
| LKW to Arrival<sup>e</sup> | 284 (151-571) | 330 (128-645) |
| Arrival to Neurology Evaluation<sup>f</sup> | 7 (3-13) | 13 (7-21) |
| Arrival to CT<sup>g</sup> | 8 (3-13) | 13 (6-26) |
| LKW to CT | 302 (168-628) | 376 (148-700) |
| LKW to CT $\leq 4.5$ hours, n (%) | 62 (41%) | 62 (41%) |
| LKW to CT $\leq 6$ hours, n (%) | 85 (57%) | 72 (48%) |
| LKW to tPA Initiation | 136 (106-178) | 157 (105-181) |
| Arrival to OR for Thrombectomy | 63 (53-79) | N/A |
| LKW to Thrombectomy Reperfusion | 368 (247-708) | N/A |

a) “Other or Unknown” race includes Asian, Black and Native Hawaiian, Filipino, unspecified other race, or not recorded. b, c, e-g) Characteristic was not documented for the following numbers of patients: b) 9, c) 23, e) 15, f) 41, and g) 15. d) CT refers to imaging performed at this facility; some patients received tPA at outside hospital prior to arrival.

### Model Performance

The Inception module-based network was selected at a threshold of 0.390 for LVO present. Results for cross-validation test sets across 10 folds as mean±95% CI are AUPRC 0.871±0.094, ROC-AUC 0.920±0.051, accuracy 85.8±4.7%, sensitivity 94.3±6.0%, precision 80.9±8.7%, specificity 79.0±9.7%, and F1 score 0.863±0.051. Results for the external validation set evaluated on each fold’s model are AUPRC 0.742±0.018, ROC-AUC 0.852±0.004, accuracy 81.2±2.7%, sensitivity 96.7±3.4%, precision 74.2±3.3%, specificity 65.7±6.8%, and F1 score 0.838±0.019.

### Characterization and Error Analysis

At the patient level, averaged results correspond to 145 TP, 5 FN, 39 FP, and 111 TN. LVO was identified for all 32 ICA occlusions, 78 (95%) M1 occlusions, 35 (97%) M2 occlusions, and 82 (99%) patients who had subsequent endovascular thrombectomy. Performance was similar for last known well (LKW) to CT (includes NCCT and CTA) interval in early (within 6 hours) and late (greater than 6 hours) thrombectomy time windows, with accuracy of 86% and 85%, respectively.

Table 2 describes TP versus FN and FP versus TN comparisons. Statistically significant differences were present for the following characteristics: history of smoking in 5 (100%) FN versus 48 (33%) TP (P=0.005), receipt of tPA in 16 (41%) FP versus 22 (20%) TN (P=0.017), and receipt of tPA specifically among patients with LKW to CT interval within 4.5 hours in 12 (71%) FP versus 18 (40%) TN (P=0.046). Image intensity window settings were lower for misclassified examples, with window level median (IQR) of 160 (130-200) for FP versus 180 (150-200) for TN (P=0.026), width 480 (310-660) for FP versus 600 (460-720) for TN (P=0.004), and width 580 (440-740) for TP versus 380 (380-400) for FN (P=0.016); window level 170 (150-210) for TP versus 150 (150-150) for FN was not significant (P=0.150). Aggregated for incorrect versus correct predictions, window level median (IQR) is 150 (130-185) versus 180 (150-202) (P=0.019) and width is 440 (320-595) versus 600 (460-725) (P<0.001).

**Table 2:**
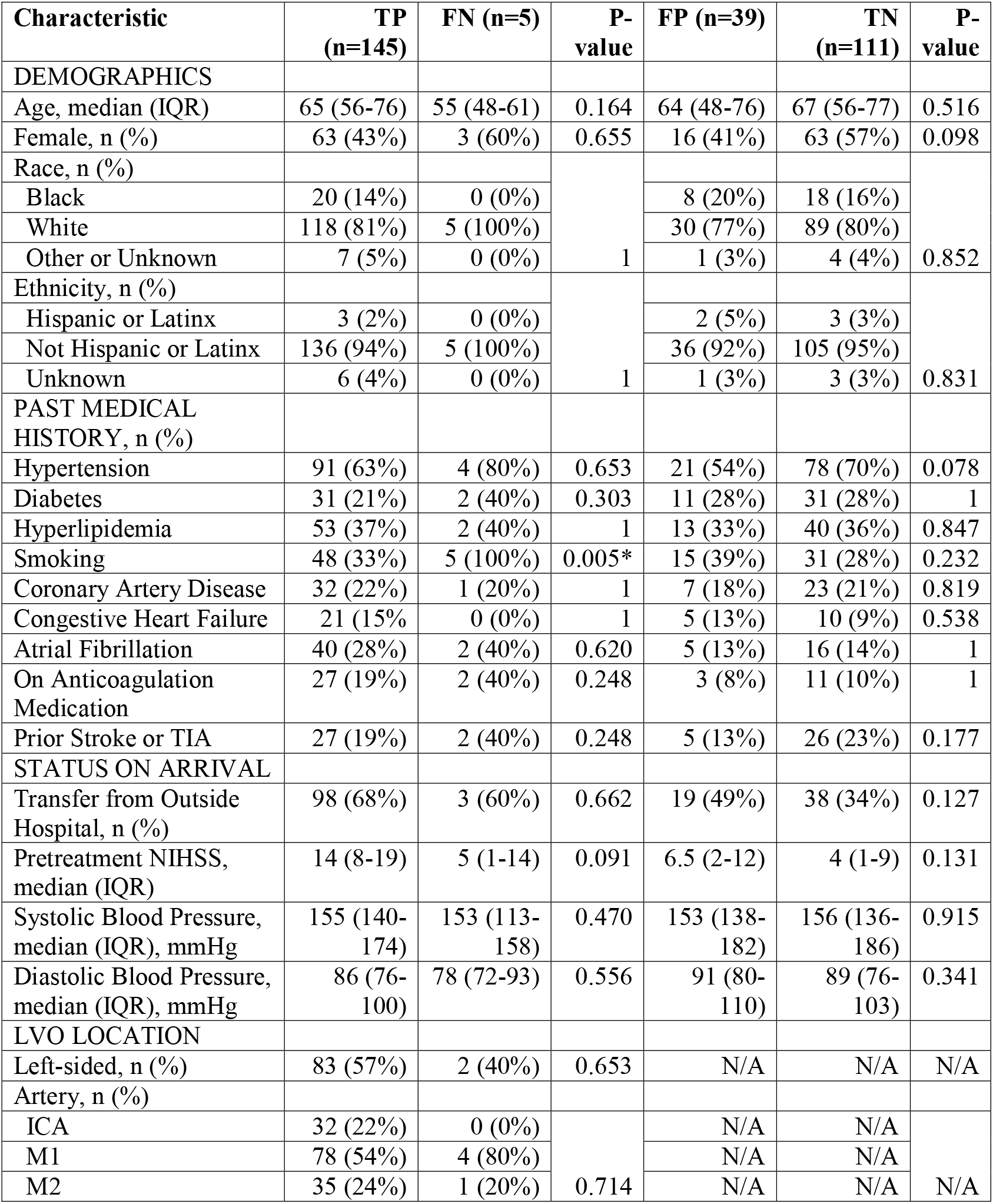
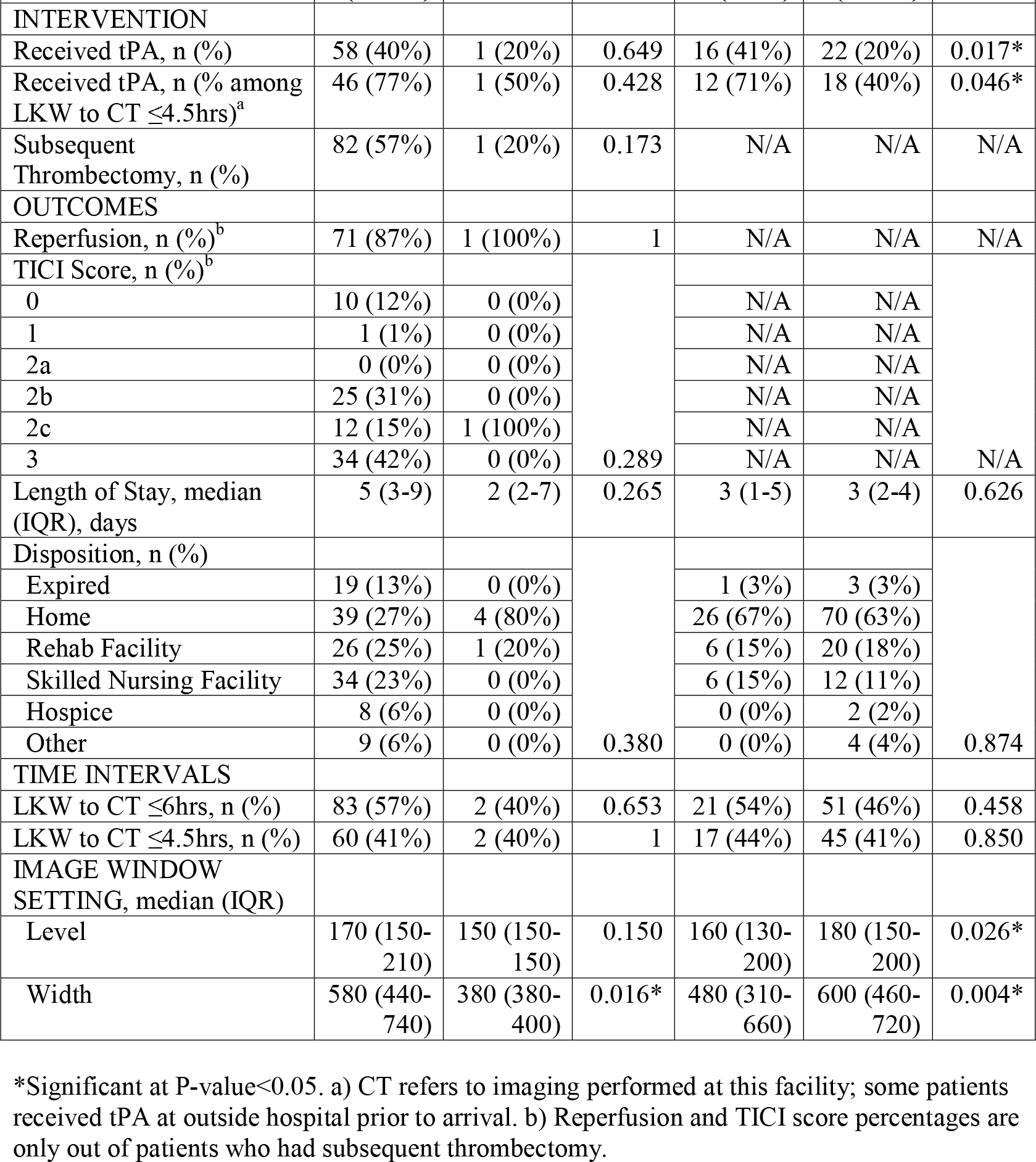
Comparison of TP versus FN and FP versus TN patient and image characteristics

| Characteristic | TP<br>(n=145) | FN (n=5) | P-value | FP (n=39) | TN<br>(n=111) | P-value |
| --- | --- | --- | --- | --- | --- | --- |
| DEMOGRAPHICS |  |  |  |  |  |  |
| Age, median (IQR) | 65 (56-76) | 55 (48-61) | 0.164 | 64 (48-76) | 67 (56-77) | 0.516 |
| Female, n (%) | 63 (43%) | 3 (60%) | 0.655 | 16 (41%) | 63 (57%) | 0.098 |
| Race, n (%) |  |  | 1 |  |  | 0.852 |
| Black | 20 (14%) | 0 (0%) |  | 8 (20%) | 18 (16%) |  |
| White | 118 (81%) | 5 (100%) |  | 30 (77%) | 89 (80%) |  |
| Other or Unknown | 7 (5%) | 0 (0%) |  | 1 (3%) | 4 (4%) |  |
| Ethnicity, n (%) |  |  | 1 |  |  | 0.831 |
| Hispanic or Latinx | 3 (2%) | 0 (0%) |  | 2 (5%) | 3 (3%) |  |
| Not Hispanic or Latinx | 136 (94%) | 5 (100%) |  | 36 (92%) | 105 (95%) |  |
| Unknown | 6 (4%) | 0 (0%) |  | 1 (3%) | 3 (3%) |  |
| PAST MEDICAL HISTORY, n (%) |  |  |  |  |  |  |
| Hypertension | 91 (63%) | 4 (80%) | 0.653 | 21 (54%) | 78 (70%) | 0.078 |
| Diabetes | 31 (21%) | 2 (40%) | 0.303 | 11 (28%) | 31 (28%) | 1 |
| Hyperlipidemia | 53 (37%) | 2 (40%) | 1 | 13 (33%) | 40 (36%) | 0.847 |
| Smoking | 48 (33%) | 5 (100%) | 0.005* | 15 (39%) | 31 (28%) | 0.232 |
| Coronary Artery Disease | 32 (22%) | 1 (20%) | 1 | 7 (18%) | 23 (21%) | 0.819 |
| Congestive Heart Failure | 21 (15%) | 0 (0%) | 1 | 5 (13%) | 10 (9%) | 0.538 |
| Atrial Fibrillation | 40 (28%) | 2 (40%) | 0.620 | 5 (13%) | 16 (14%) | 1 |
| On Anticoagulation Medication | 27 (19%) | 2 (40%) | 0.248 | 3 (8%) | 11 (10%) | 1 |
| Prior Stroke or TIA | 27 (19%) | 2 (40%) | 0.248 | 5 (13%) | 26 (23%) | 0.177 |
| STATUS ON ARRIVAL |  |  |  |  |  |  |
| Transfer from Outside Hospital, n (%) | 98 (68%) | 3 (60%) | 0.662 | 19 (49%) | 38 (34%) | 0.127 |
| Pretreatment NIHSS, median (IQR) | 14 (8-19) | 5 (1-14) | 0.091 | 6.5 (2-12) | 4 (1-9) | 0.131 |
| Systolic Blood Pressure, median (IQR), mmHg | 155 (140-174) | 153 (113-158) | 0.470 | 153 (138-182) | 156 (136-186) | 0.915 |
| Diastolic Blood Pressure, median (IQR), mmHg | 86 (76-100) | 78 (72-93) | 0.556 | 91 (80-110) | 89 (76-103) | 0.341 |
| LVO LOCATION |  |  |  |  |  |  |
| Left-sided, n (%) | 83 (57%) | 2 (40%) | 0.653 | N/A | N/A | N/A |
| Artery, n (%) |  |  |  |  |  |  |
| ICA | 32 (22%) | 0 (0%) | 0.714 | N/A | N/A | N/A |
| M1 | 78 (54%) | 4 (80%) |  | N/A | N/A |  |
| M2 | 35 (24%) | 1 (20%) |  | N/A | N/A |  |

**Table 2: Comparison of TP versus FN and FP versus TN patient and image characteristics (continued)**
| Characteristic | TP<br>(n=145) | FN (n=5) | P-value | FP<br>(n=39) | TN<br>(n=111) | P-value |
| --- | --- | --- | --- | --- | --- | --- |
| INTERVENTION |  |  |  |  |  |  |
| Received tPA, n (%) | 58 (40%) | 1 (20%) | 0.649 | 16 (41%) | 22 (20%) | 0.017* |
| Received tPA, n (%) among LKW to CT ≤ 4.5hrs<sup>a</sup> | 46 (77%) | 1 (50%) | 0.428 | 12 (71%) | 18 (40%) | 0.046* |
| Subsequent Thrombectomy, n (%) | 82 (57%) | 1 (20%) | 0.173 | N/A | N/A | N/A |
| OUTCOMES |  |  |  |  |  |  |
| Reperfusion, n (%)<sup>b</sup> | 71 (87%) | 1 (100%) | 1 | N/A | N/A | N/A |
| TICI Score, n (%)<sup>b</sup> |  |  | 0.289 |  |  | N/A |
| 0 | 10 (12%) | 0 (0%) |  | N/A | N/A |  |
| 1 | 1 (1%) | 0 (0%) |  | N/A | N/A |  |
| 2a | 0 (0%) | 0 (0%) |  | N/A | N/A |  |
| 2b | 25 (31%) | 0 (0%) |  | N/A | N/A |  |
| 2c | 12 (15%) | 1 (100%) |  | N/A | N/A |  |
| 3 | 34 (42%) | 0 (0%) |  | N/A | N/A |  |
| Length of Stay, median (IQR), days | 5 (3-9) | 2 (2-7) | 0.265 | 3 (1-5) | 3 (2-4) | 0.626 |
| Disposition, n (%) |  |  | 0.380 |  |  | 0.874 |
| Expired | 19 (13%) | 0 (0%) |  | 1 (3%) | 3 (3%) |  |
| Home | 39 (27%) | 4 (80%) |  | 26 (67%) | 70 (63%) |  |
| Rehab Facility | 26 (25%) | 1 (20%) |  | 6 (15%) | 20 (18%) |  |
| Skilled Nursing Facility | 34 (23%) | 0 (0%) |  | 6 (15%) | 12 (11%) |  |
| Hospice | 8 (6%) | 0 (0%) |  | 0 (0%) | 2 (2%) |  |
| Other | 9 (6%) | 0 (0%) |  | 0 (0%) | 4 (4%) |  |
| TIME INTERVALS |  |  |  |  |  |  |
| LKW to CT ≤ 6hrs, n (%) | 83 (57%) | 2 (40%) | 0.653 | 21 (54%) | 51 (46%) | 0.458 |
| LKW to CT ≤ 4.5hrs, n (%) | 60 (41%) | 2 (40%) | 1 | 17 (44%) | 45 (41%) | 0.850 |
| IMAGE WINDOW SETTING, median (IQR) |  |  |  |  |  |  |
| Level | 170 (150-210) | 150 (150-150) | 0.150 | 160 (130-200) | 180 (150-200) | 0.026* |
| Width | 580 (440-740) | 380 (380-400) | 0.016* | 480 (310-660) | 600 (460-720) | 0.004* |

\*Significant at P-value<0.05. a) CT refers to imaging performed at this facility; some patients received tPA at outside hospital prior to arrival. b) Reperfusion and TICI score percentages are only out of patients who had subsequent thrombectomy.

Example processed CTA MIPs of TP, FN, FP, and TN classifications are shown in Figure 2. The five FN patients had: 1) M1 occlusion and underwent successful thrombectomy with thrombolysis in cerebral infarction (TICI) 2c reperfusion, 2) M2 occlusion yet not a candidate for thrombectomy due to good collateral flow and NIHSS of 1, 3) M1 occlusion with likely chronic vasculopathy due to polysubstance use, and 4) and 5) M1 occlusion felt to be chronic and not amenable to intervention in these patients who both had known prior stroke. Among 39 FP, only 11 patients had no pathology or artifacts noted. Fifteen FP had distal non-LVO occlusion: six in the distal M2, six in the third segment (M3) of the middle cerebral artery, one unspecified M2 or M3 short segment sylvian middle cerebral artery thrombosis, one likely chronic cervical ICA occlusion that was documented and verified as not LVO, and one with multifocal occlusions in the anterior cerebral, vertebral, and posterior cerebral arteries. Seven FP had severe unifocal or multifocal stenosis noted. Five FP had other pathology not necessarily specific to CTA findings: basilar artery focal filling defect, lacunar infarct of unknown chronicity, remote cerebellar infarct, likely thrombosed vertebral aneurysm plus cerebellar and thalamic strokes, and decreased mean transit time and cerebral blood flow in frontoparietal M3 region found on subsequent perfusion imaging without CT correlate. Three FP had artifact specified in the radiology report but were still interpretable as far as LVO label and thus had not been excluded from the dataset: 1) mild motion artifact in one with M3 occlusion, 2) limited by venous contamination in one with lacunar infarct, and 3) suboptimal contrast opacification and quantum mottle, streak, and motion artifact.

**Figure 2:**
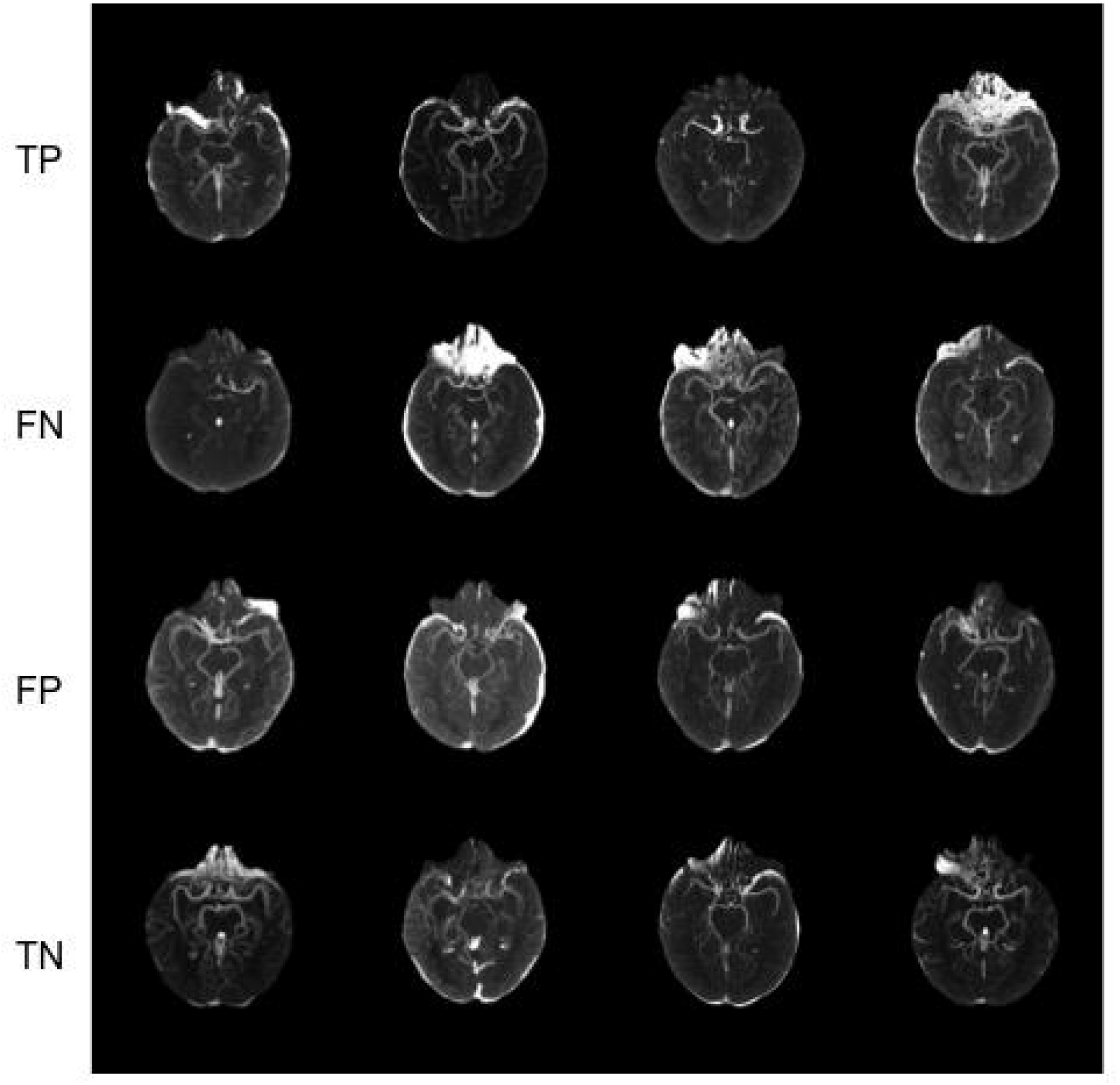
Example processed CTA MIPs of TP, FN, FP, and TN classifications. From left to right, TP in first row: 1) left ICA occlusion with window width (W) 480 and level (L) 150, 2) right M1 occlusion, W1000 L250, 3) left M1 occlusion, W600 L180, 4) right M2 occlusion, W400 L140. FN in second row: 1) right M1 with subsequent thrombectomy, W400 L150, 2) chronic left M1 occlusion and prior strokes, W420 L150, 3) left M2 occlusion with good collaterals and NIHSS 1, W380 L150, 4) chronic right M1 occlusion and prior strokes, W380 L150. FP in third row: 1) venous contamination and lacunar infarct, W320 L130, 2) no noted pathology, W240 L110, 3) right M3 occlusion, W420 L150, 4) multifocal stenosis, W480 L160. TN in fourth row: 1) W820 L 220, 2) W400 L140, 3) W520 L170, 4) W800 L230.

## Conclusions

The LVO detection tool developed has high accuracy and sensitivity that holds true across patient characteristics with few exceptions. Analysis revealed statistically significant differences for the following variables: lower window settings for misclassifications, smoking history more frequent in FN than TP, and tPA administered to more FP than TN. Investigation of misclassifications revealed that many FP had pathology warranting detection including distal occlusions, and four of five FN had chronic changes or other factors that precluded them from being candidates for endovascular thrombectomy.

Our model cross-validation performance with AUPRC 0.87, ROC-AUC 0.92, accuracy 86%, sensitivity 94%, specificity 79%, and precision 81% is comparable to prior studies of automated LVO detection and falls slightly short of radiologist performance of accuracy 96%, sensitivity 96%, and specificity 97% in a study by Boyd et al.^43^ Table 3 compares our results with prior work on automated LVO detection from CTA, including studies using the commercial platforms Viz.ai and RapidLVO as well as two studies using non-commercial models.^12–17^ LVO definitions were similar to our study with the additional inclusion of anterior cerebral artery occlusions by Sheth et al. and posterior circulation occlusions by Stib et al. Some of these studies trained models on larger datasets and/or included multiple institutions and CT scanners, which may inherently improve generalizability, but this cannot be assumed. Several reported patient characteristics of their study population without subsequent analysis of results in the context of these features. The only exception is the study by Dehkharghani et al. using RapidLVO software, which demonstrated similar performance across age groups, sex, location within or outside the United States, and CT scanner manufacturers.^44^ Our study found similar results with age and sex in addition to other demographics. With only five FN, it is unclear how much emphasis to place on the finding of higher prevalence of smoking history compared to TP, yet this factor may underlie the chronic vasculature changes found in three FN.^21^ Our model performed consistently across the artery of occlusion (100% ICA, 95% M1, and 97% M2), whereas a previous study found that the sensitivity of LVO detection using RapidLVO software was higher for ICA or M1 than M2 occlusions (97% versus 90%) for their dataset.^15^ Our model also did not perform differently for patients presenting within 6 hours of stroke onset versus later than 6 hours; current guidelines recommend additional perfusion imaging for patients presenting later than 6 hours from LKW, yet this advanced imaging is less available at many sites, and its necessity is under scrutiny.^1,45,46^ Furthermore, LVO was identified by our model in all but one patient who ultimately had thrombectomy, supporting its potential use as a tool in deciding which patients require transport to thrombectomy-capable centers.

**Table 3:** Comparison with prior studies on automated LVO detection from CTA. Cross-validation test set (CV) and external validation set (EV) mean results are listed for this study.

| **Study** | **Software/CNN** | **# Patients** | **ROC-AUC** | **Sensitivity** | **Specificity** |
| --- | --- | --- | --- | --- | --- |
| This study | Inception V1 module-based | 300 | 0.92 CV, 0.85 EV | 0.94 CV, 0.97 EV | 0.79 CV, 0.66 EV |
| Barreira et al.<sup>12</sup> | Viz.ai | 875 | 0.86 | 0.90 | 0.83 |
| Chatterjee et al.<sup>13</sup> | Viz.ai | 650 | - | 0.82 | 0.94 |
| Amukotuwa et al., single-center<sup>14</sup> | RapidLVO | 477 | 0.86 | 0.97 | 0.81 |
| Amukotuwa et al., multicenter<sup>15</sup> | RapidLVO | 926 | 0.95 | 0.95 | 0.79 |
| Dehkharghani et al.<sup>44</sup> | RapidLVO | 217 | 0.99 | 0.96 | 0.98 |
| Sheth et al.<sup>16</sup> | DeepSymNet (novel CNN) | 297 | 0.88 | - | - |
| Stib et al.<sup>17</sup>: single-phase CTA | DenseNet-121 | 540 | 0.74 | 0.77 | 0.71 |
| Stib et al.<sup>17</sup>: multi-phase CTA | DenseNet-121 | 540 | 0.89 | 1 | 0.77 |

Distal M2 or further distal occlusions were not labeled as LVO for the purposes of this study and had been documented as too distal for retrieval by thrombectomy. The highest level of recommendation for thrombectomy eligibility includes only ICA and M1 occlusion, while benefits are uncertain for M2, M3, or other occlusions.^1^ Occlusions that are harder to access directly are nonetheless important to identify and treat early with intravenous tPA, and there is evidence of benefit of emerging thrombectomy techniques and intra-arterial tPA for distal occlusions.^47–50^ Thus, it may in fact be advantageous that our model classified as positive the fifteen FP with distal occlusion and additional five with thrombosis or other findings indicative of infarct. This pathology may be related to the higher frequency among FP than TN of administration of intravenous tPA, which was not attributable to any difference in time of presentation. Stib et al. discussed one FP for which they later found an infarct based on magnetic resonance imaging (MRI) and raised the idea that their model may be able to detect smaller infarcts than even radiologists can see on CTA.^17^ We did not investigate MRI findings, but it is possible that a similar idea applies in our study to one FP with CT perfusion findings indicating potential injury to the M3 region and others with signs of infarct on NCCT. Stenosis was our other prevalent finding among FP and is common in other studies, including 16 of 31 FP with intracranial atherosclerosis in the study by Chatterjee et al.^13^ RapidLVO software relies on detection of asymmetric vasculature density, and Amukotuwa et al. found that FP commonly had natural inter-hemispheric variation in vasculature, ipsilateral decreased vascular density due to stenosis or prior infarct, or contralateral increased vascular density due to opacified venous structures, aneurysm, hematoma, or reactive hyperemia.^14,15^ One FP from our study had noted venous contamination that may have contributed to misclassification. The slice range for MIP generation was selected based on a sample to optimize visualization of anterior circulation but may not have captured the entire anterior circulation for all CTAs due to anatomic variation, which may have created a false appearance of vascular asymmetry or cutoffs in some FP.

Amukotuwa et al. found FN to commonly have short-segment or incomplete occlusion with distal reconstitution, small nondominant branch occlusions, or collaterals resulting in normal or increased vessel density on the side of the occlusion.^14,15^ One FN from our study had an acute LVO with robust collaterals, and several other FN with chronic occlusions or vasculopathy may also have had time for collateral formation. Occlusions with robust collaterals have slower infarct progression and thus lower consequence of delayed identification and treatment. Only one patient in our study with acute LVO who was a thrombectomy candidate was not identified, which may be explained by lower window settings and likely poorer image quality among FN.

The windowing findings highlight the importance of considering input data quality when using an automated algorithm for clinical decision support. Windowing was adjusted to optimize vessel visualization for each CTA, and lower settings among misclassifications indicate lower signal-to-noise ratio likely due to poor contrast attenuation. Contrast enhancement is variable due to patient factors including weight and cardiac output, contrast factors such as injection duration and rate, and scan factors including duration and delay.^24^ While target contrast and scan parameters exist for each type of CTA scan, patient variation is difficult to control for. Ability to detect LVO by visual inspection is the primary consideration for whether CTA quality is sufficient for both the acute stroke triage setting and inclusion criteria for this study. Thus, even the three FP with documented limitation by artifacts were included, as well as scans for which it was relatively difficult to achieve similar optimization of vessel visualization as other scans, which were often those requiring a lower window range. Optimized window settings for CTA have been found to 1) reduce variability in vessel measurement relative to variation in contrast attenuation and 2) improve accuracy of CTA source image-based ASPECTS regardless of rater experience or specialty.^51,52^ Prior LVO detection studies have not discussed windowing, and some used vessel segmentation or tubular filtering methods that enhance signal-to-noise ratio and are perhaps less dependent on contrast attenuation differences or other factors underlying image variation.^15,17^ It is possible that if our algorithm was trained on a larger dataset, it could identify LVO despite poor contrast attenuation or artifact, and this may also obviate the need for individualized window adjustment beforehand. However, it is also possible that the effect of CTA quality of individual images has been overlooked in studies using different methods and often larger datasets. Scans with poorer quality should not simply be excluded in algorithm training or application, as this would limit the fundamental utility and applicability as a tool for triage. Chatterjee et al. excluded cases with inadequate contrast and motion artifact in their study yet mentioned ongoing efforts to optimize detection in poor quality CTAs.^13^

Manual chart review by one author is a possible source of error. Posterior circulation and anterior cerebral artery occlusions were excluded due to rarity in this dataset, yet these are included in some LVO definitions because carefully selected patients may be candidates for thrombectomy.^1^ Image processing included manual steps of individual image window range adjustment and selection based on visual inspection of the axial slice range used to generate the MIP. This limits utility of the tool in clinical practice without further development. Windowing based on visual inspection is a potential source of inconsistency, although it was performed by the same author for all images which provides some consistency. Note that during development, the model was not able to learn when the same window range was applied to all images, so individual adjustment was a necessary processing step; with sufficient data, windowing may be another feature that can be learned. The MIP slice range selection is a potential source of error as previously discussed, yet the reduction from three-dimensional to two-dimensional data enabled the model to learn effectively and efficiently from a small dataset of 300 images. Model training, performance, characterization analysis, and generalizability are limited by this being a single institution study of 300 patients. The small study size is the major limitation in training complex deep learning algorithms and the consequent performance and robustness across features such as diverse anatomy and image quality. Although the algorithm could identify LVO with high accuracy and sensitivity and without performance bias based on demographic factors and most stroke risk factors within our own study population, the applicability in a larger context is biased by the patients included.^18,19^

This study is the first to characterize performance of a machine learning based LVO detection tool in the context of comprehensive patient and imaging characteristics, with key findings of lower window settings among misclassifications and distal occlusion or stenosis prevalent among false positives. Image quality should be carefully considered when using automated LVO detection for decision support. The tool is otherwise highly accurate and sensitive across patient characteristics, demonstrating broad applicability that supports the use of machine learning to aid stroke triage.

## Data Availability

All data produced in the present study are available upon reasonable request to the authors.

## Acknowledgements

We are grateful to Kiersten Espaillat, Stroke Program Manager at Vanderbilt University Medical Center, for assistance in identifying patients for this study and sharing the stroke alert database. We are also grateful to MASI lab members Riqiang Gao, Cailey Kerley, Karthik Ramadass, and Praitayini Kanakaraj for their assistance.

